## Supplementary Materials for "Care seeking for diarrheal illness: a systematic review and meta-analysis"

Table of Contents


**Supplementary Methods** **2**

Systematic review search methods 2

**Supplementary Figures** **5**

Figure S1. Data coverage by geography 5

Figure S2. Data coverage over time 6

Figure S3. Proportion that sought care at a hospital or clinic by region 7

Figure S4. Proportion that sought care at a hospital or clinic by income group 8

Figure S5. Proportion that sought care by study quality in LMICs 9

Figure S6. Proportion that sought care by source and income group in LMICs 10

Figure S7. Proportion that sought care by source and case definition in LMICs 11

Figure S8. Proportion that sought care by source and observation characteristic for clinic-based studies 12

Figure S9. Forest plot of care seeking at a hospital or clinic in LMICs 13

Figure S10. Forest plot of hypothetical care seeking at a hospital or clinic in LMICs 16

Figure S11. Forest plot of hypothetical care seeking at a hospital or clinic in HICs 17

Figure S12. Forest plot of care seeking for general diarrhea in LMICs 18

Figure S13. Forest plot of care seeking for severe diarrhea or cholera in LMICs 21

Figure S14. Forest plot of care seeking for gastroenteritis or other etiologies in LMICs 21

**Supplementary Tables** **22**

Table S1. Standardized categories for sources of care and examples from studies 22

Table S2. Univariate analyses of factors associated with variation in care seeking 23

Table S3. Tests for potential confounding with severe diarrhea or cholera case definition 24

Table S4. Sub-analyses of associations with outbreak context and recall period 25

### Supplementary Methods

#### Systematic review search methods

**Search methods for identification of studies:**
To identify studies to include or consider for this systematic review, the review team worked with a medical librarian (AGS) to develop detailed search strategies for each database. The PRISMA-S extension was followed for search reporting. The medical librarian (AGS) developed the search for PubMed (NLM) and translated the search for every database searched. The PubMed (NLM) search strategy was reviewed by the research team to check for accuracy and term relevancy, and all final searches were peer-reviewed by another medical librarian following the PRESS checklist. The databases included in this search are PubMed (NLM), Embase (Elsevier, embase.com), Web of Science (Clarivate Analytics), and Global Index Medicus (World Health Organization), which were searched using a combination of keywords and subject headings. Searches were limited to articles published since 2000, in line with the review team's body of literature. All final searches were performed on January 27, 2023; searches were then updated on September 3, 2024. The full search strategies as reported by the librarian are provided below.

**Search strings:**

PubMed (NLM)

| |
|---|
| (diarrhea*[tiab] OR diarrhoea*[tiab] OR cholera[tiab] OR diarrhea[mesh] OR cholera[mesh]) AND (care seeking[tiab] OR healthcare seeking[tiab] OR seeking healthcare[tiab] OR seek* care[tiab] OR sought care[tiab] OR health seeking[tiab] OR health care utilization[tiab] OR health care utilisation[tiab] OR healthcare utilization[tiab] OR healthcare utilisation[tiab] OR health care use[tiab] OR healthcare use[tiab] OR health services utilization[tiab] OR health services utilisation[tiab] OR health services use[tiab] OR treatment seeking[tiab] OR seek* treatment[tiab] OR health behavior[tiab] OR health behaviour[tiab] OR "patient acceptance of health care"[mesh] OR "health services accessibility"[mesh] OR "health knowledge, attitudes, practice"[mesh] or "health behavior"[mesh:noexp]) AND (2000:2024[pdat]) |

Embase (Elsevier, embase.com)

| | |
|---|---|
| #1 | (diarrhea*:ab,ti or diarrhoea*:ab,ti or cholera:ab,ti or diarrhea/exp or cholera/exp) |
| #2 | (‘care seeking’:ab,ti or ‘healthcare seeking’:ab,ti or (seeking NEAR/3 healthcare):ab,ti or (seek* NEAR/3 care):ab,ti or (sought NEAR/3 care):ab,ti or ‘health seeking’:ab,ti or ‘health care utilization’:ab,ti or ‘health care utilisation’:ab,ti or ‘healthcare utilization’:ab,ti or ‘healthcare utilisation’:ab,ti or ‘health care use’:ab,ti or ‘healthcare use’:ab,ti or ‘health services utilization’:ab,ti or ‘health services utilisation’:ab,ti or ‘health services use’:ab,ti or ‘treatment seeking’:ab,ti or (seek* NEAR/3 treatment):ab,ti or ‘health behavior’:ab,ti or ‘health behaviour’:ab,ti or ‘patient attitude’/de or ‘health care utilization’/exp or ‘help seeking behavior’/exp or ‘health behavior’/de) |
| #3 | #1 and #2 and [2000-2024]/py |

Web of Science (Clarivate Analytics)

| | |
|---|---|
| #1 | TS=(diarrhea* or diarrhoea* or cholera) |
| #2 | TS=(“care seeking” or “healthcare seeking” or (seeking NEAR/3 healthcare) or (seek* NEAR/3 care) or (sought NEAR/3 care) or “health seeking” or “health care utilization” or “health care utilisation” or “healthcare utilization” or “healthcare utilisation" or “health care use” or “healthcare use” or “health services utilization” or “health services utilisation" or “health services use” or “treatment seeking” or (seek* NEAR/3 treatment) or “health behavior” or “health behaviour”) |
| #3 | DOP=(2000/2024) |
| #4 | #1 and #2 and #3 |

Global Index Medicus (WHO)

| |
|---|
| (tw:(diarrhea* or diarrhoea* or cholera)) AND (tw:("care seeking" or "healthcare seeking" or "seeking healthcare" or "seek care" or "seeking care" or "sought care" or "health seeking" or "health care utilization" or “health care utilisation” or “healthcare utilization” or “healthcare utilisation" or “health care use” or “healthcare use” or “health services utilization” or “health services utilisation" or “health services use” or “treatment seeking” or "seek treatment" or "seeking treatment")) AND (year_cluster:[2000 TO 2024]) |

**Summary of the search results from databases:**
PubMed (NLM, 1809-present): 2372 results
Embase (Elsevier, embase.com, 1974-present): 3561 results
Web of Science (Clarivate Analytics, 1900-present): 1123 results
Global Index Medicus (World Health Organization): 13 results

The search resulted in 7069 studies and 1881 duplicate studies were found and omitted using Covidence or by manual identification. This resulted in 5188 records to screen from databases or registers.

#### Supplementary Figures

##### Figure S1. Data coverage by geography

Number of observations in the primary dataset at each administrative level as defined by GADM (https://gadm.org/) by country. Countries with >10 observations displayed as 10.

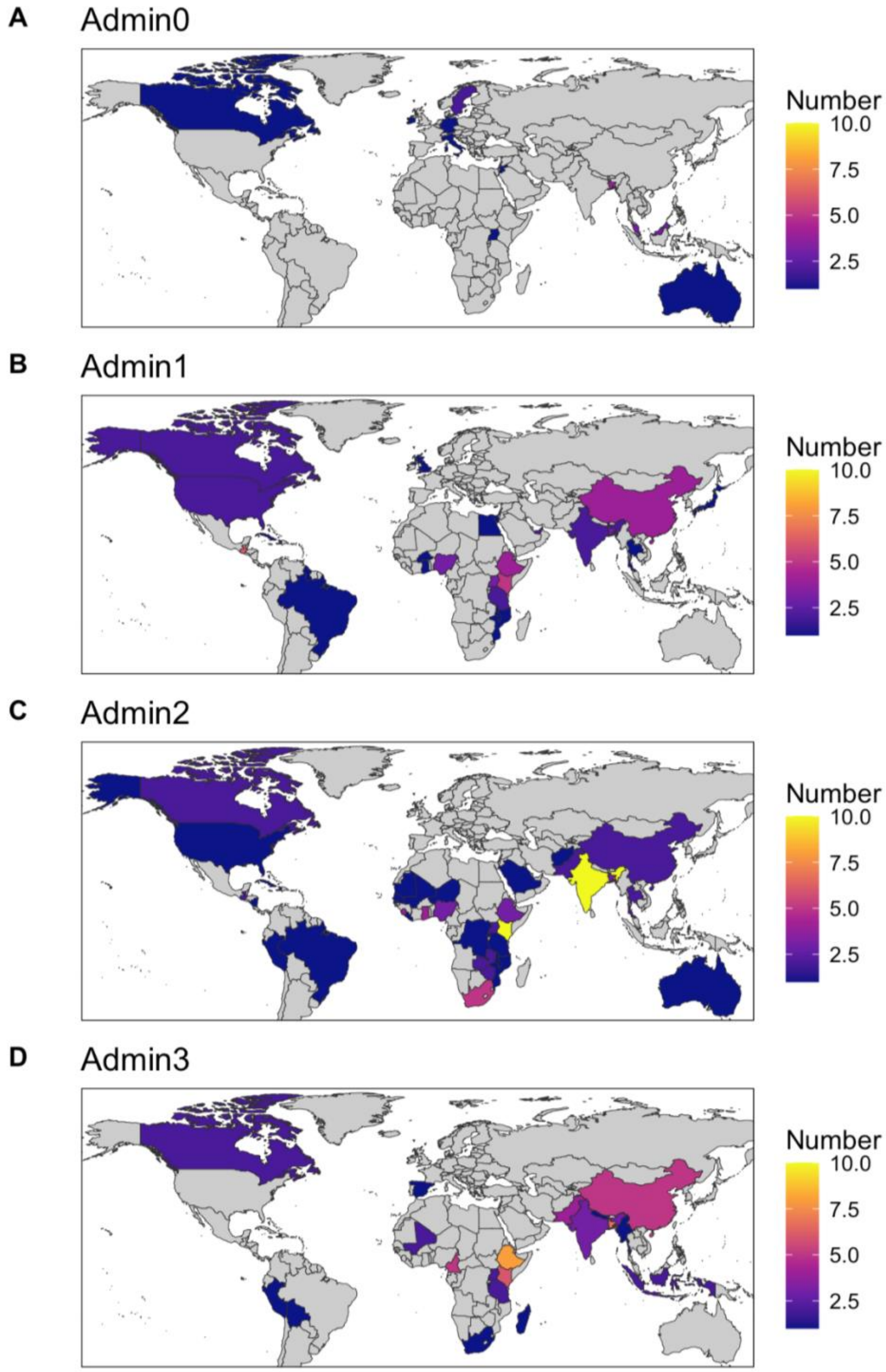

Figure S2. Data coverage over time

Number of observations in the primary dataset at each administrative level as defined by GADM (https://gadm.org/) by country within different time periods. Countries with >10 observations displayed as 10. Year represents the year sampling was completed. Excludes 6 studies missing a study end date.

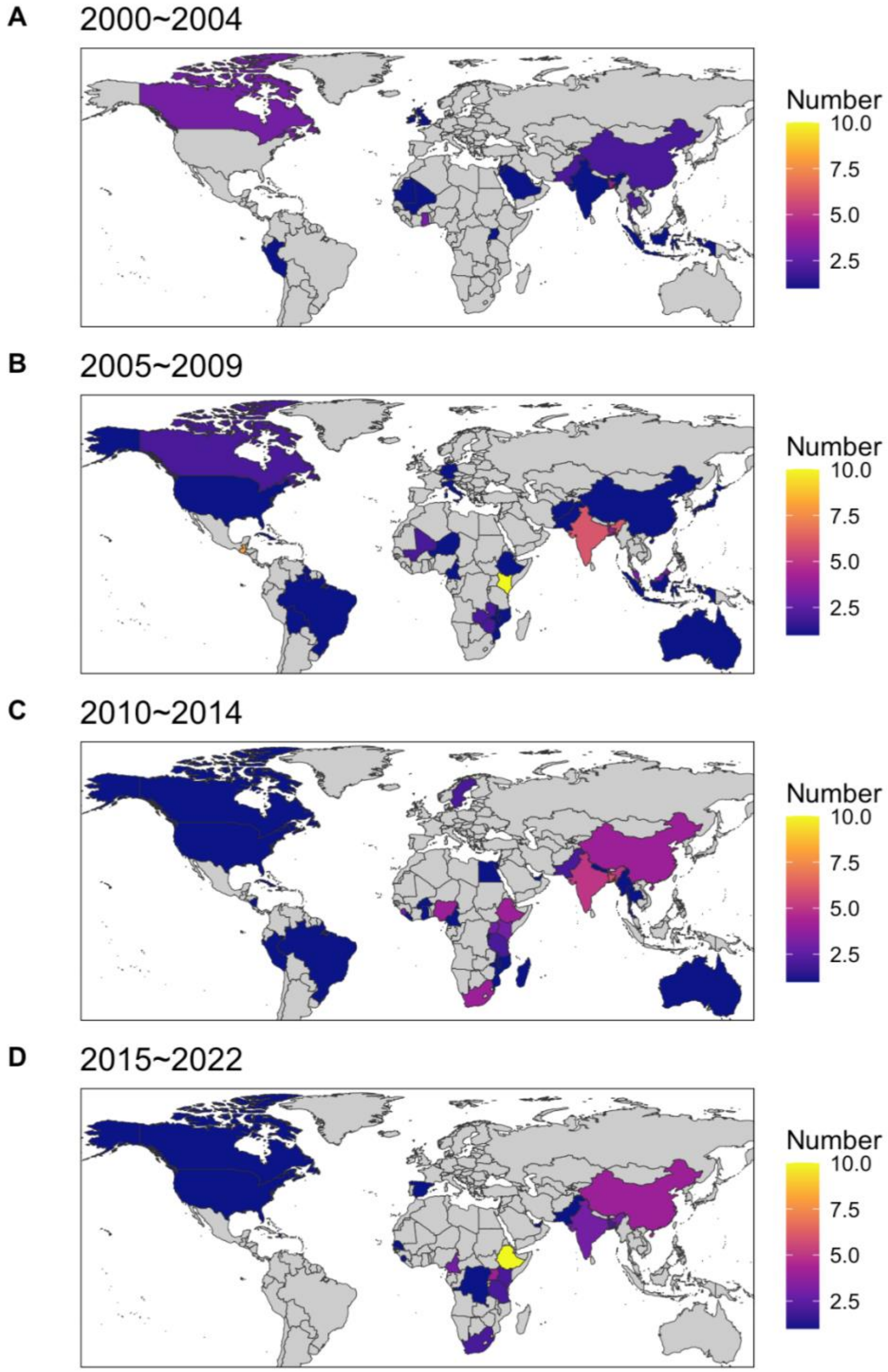

Figure S3. Proportion that sought care at a hospital or clinic by region

Proportion of respondents that reported seeking care for diarrheal illness at a hospital or clinic grouped by A) geographic region and B) geographic region and case definition. Each point represents an observation. Boxes represent the median and interquartile range (IQR) of the proportion for each group. Lines extend from the top and bottom of box to the largest proportion value no further than 1.5 * IQR from the box.

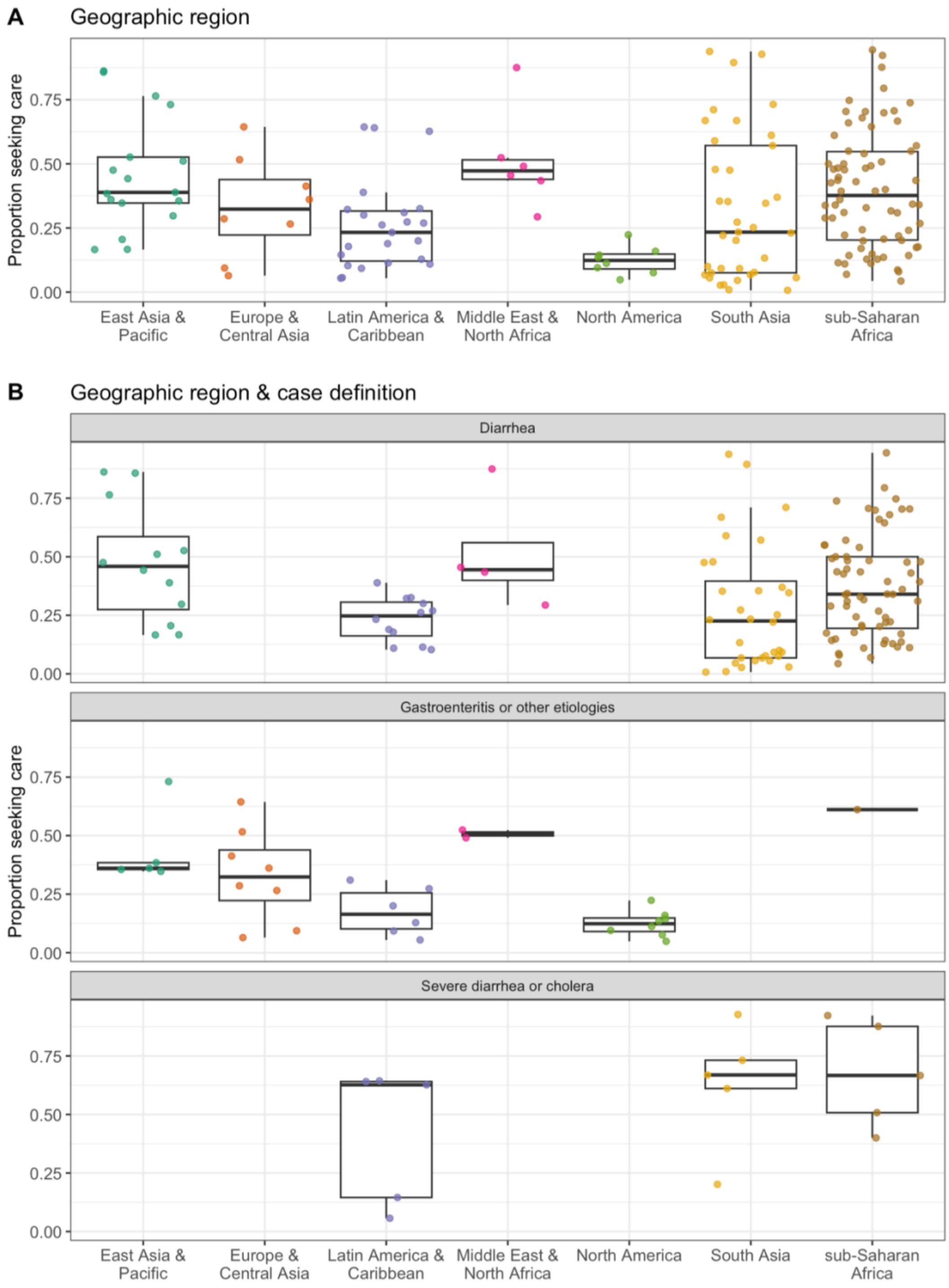

#### Figure S4. Proportion that sought care at a hospital or clinic by income group

Proportion of respondents that reported seeking care for diarrheal illness at a hospital or clinic grouped by A) World Bank income group and B) World Bank 2023-2024 income group and case definition. Each point represents an observation. Boxes represent the median and interquartile range (IQR) of the proportion for each group. Lines extend from the top and bottom of box to the largest proportion value no further than 1.5 * IQR from the box.

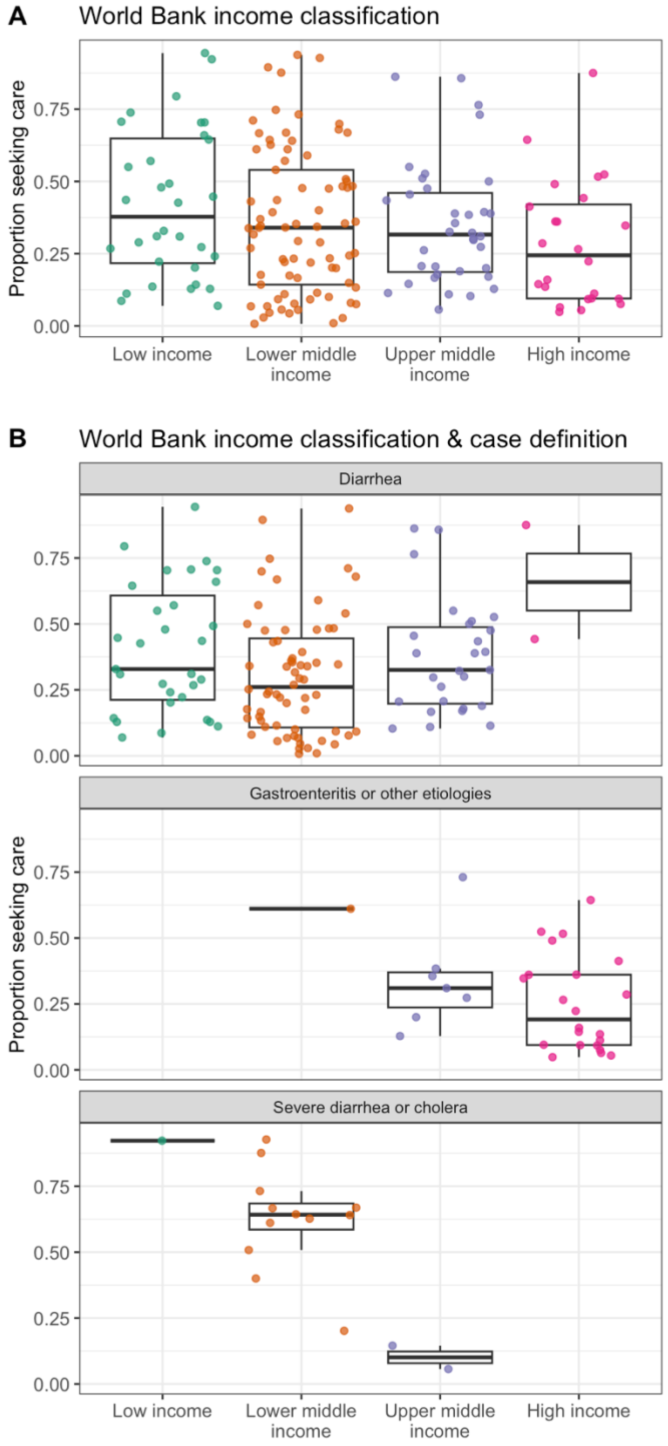

Figure S5. Proportion that sought care by study quality in LMICs

Proportion of respondents in low- and middle-income countries (LMICs) that reported seeking care for diarrheal illness at a hospital or clinic grouped by A) overall quality score, B) whether the data is internally consistent (i.e., numbers for the proportion seeking care were identical throughout the abstract, text, and tables), C) whether the manuscript included a justification for their sample size, and D) whether the non-response rate was reported. Overall quality score is the sum of B-D. Each point represents an observation. Boxes represent the median and interquartile range (IQR) of the proportion for each group. Lines extend from the top and bottom of box to the largest proportion value no further than 1.5 * IQR from the box.

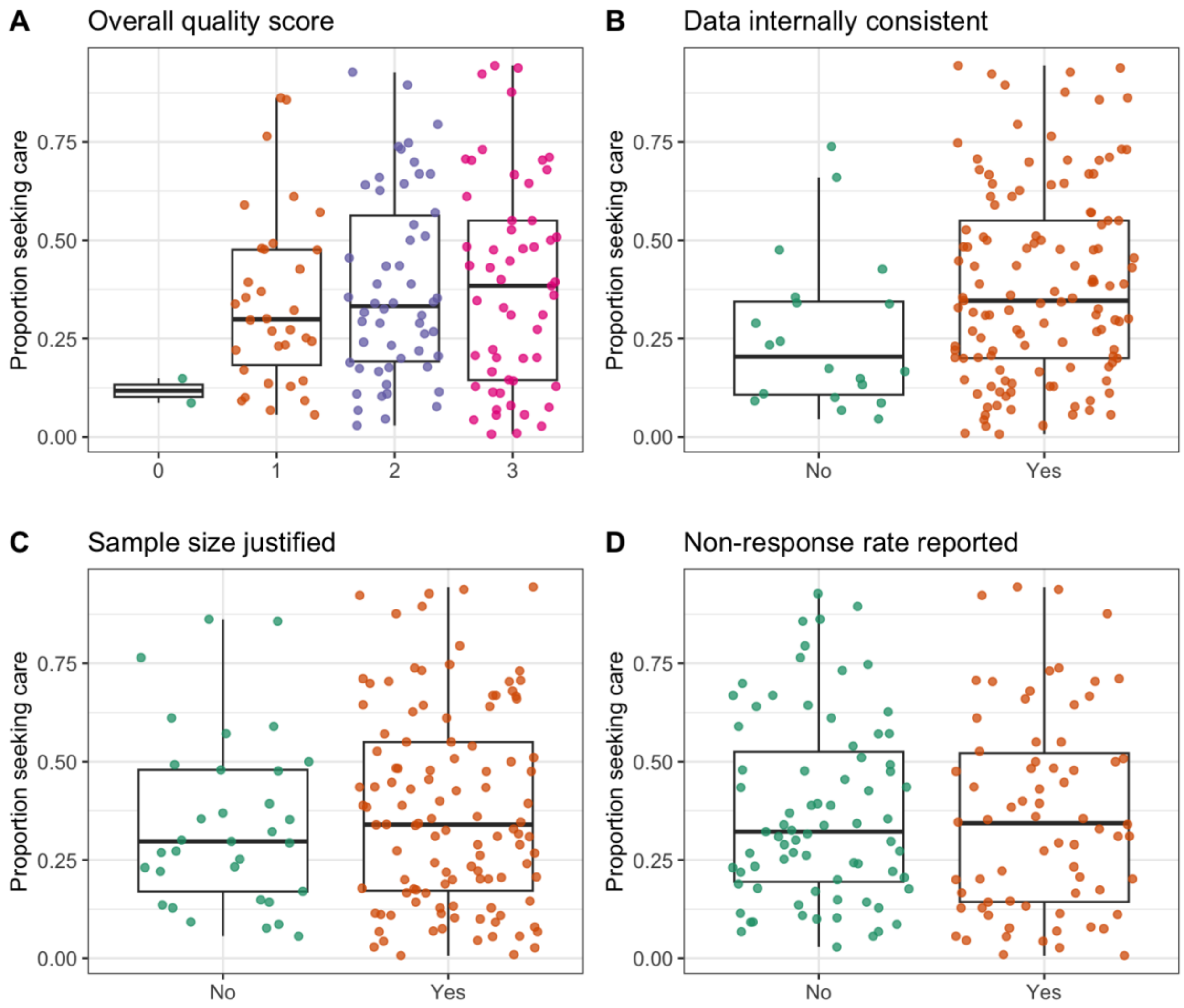

Figure S6. Proportion that sought care by source and income group in LMICs

Proportion of respondents in LMICs that reported seeking care for diarrheal illness at all categories of care sources by the World Bank income group of the country the study was conducted in. Each point represents an observation. Boxes represent the median and interquartile range (IQR) of the proportion for each group. Lines extend from the top and bottom of box to the largest proportion value no further than 1.5 * IQR from the box.

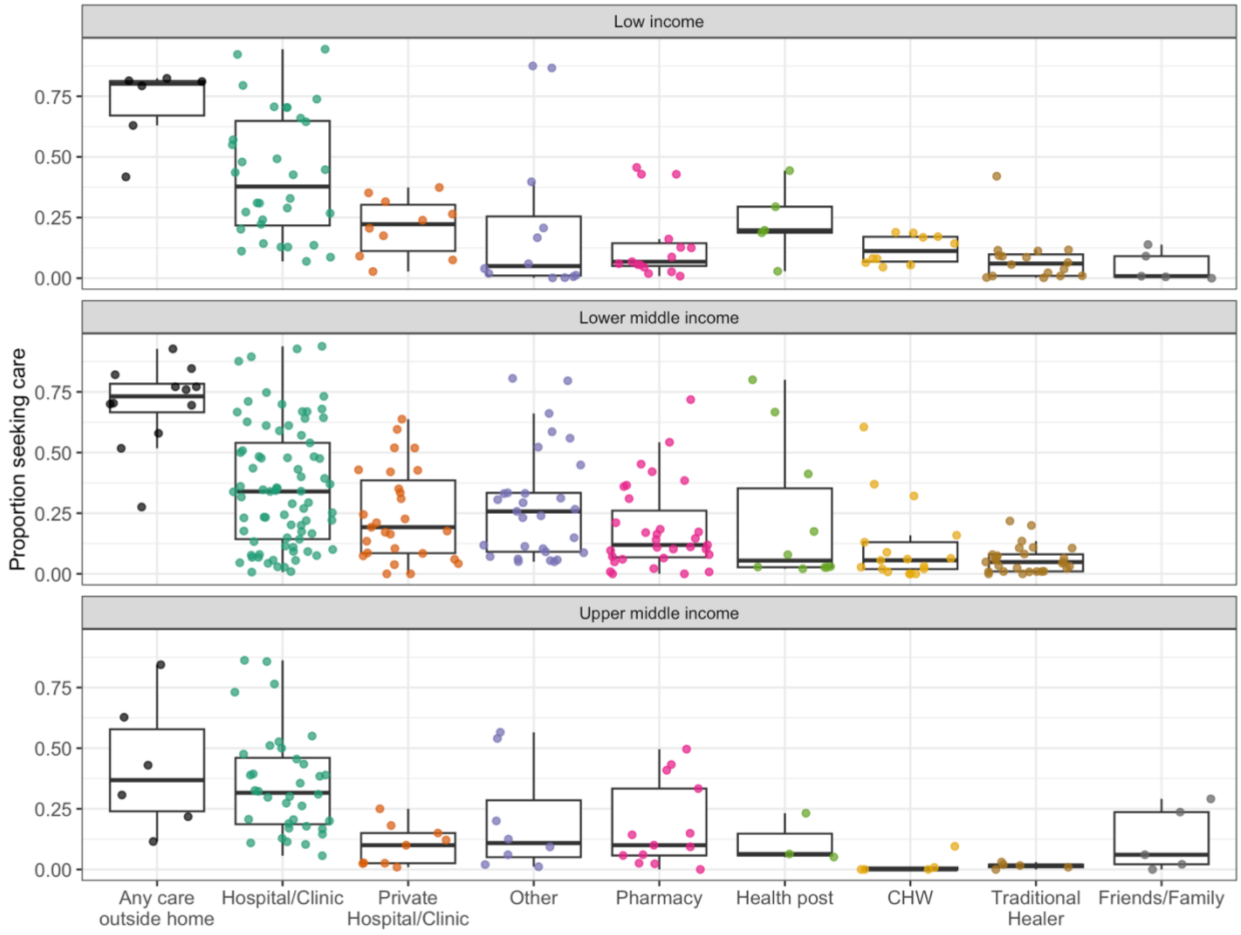

Figure S7. Proportion that sought care by source and case definition in LMICs

Proportion of respondents in LMICs that reported seeking care for diarrheal illness at all categories of care sources by diarrhea case definition. Each point represents an observation. Boxes represent the median and interquartile range (IQR) of the proportion for each group. Lines extend from the top and bottom of box to the largest proportion value no further than 1.5 * IQR from the box.

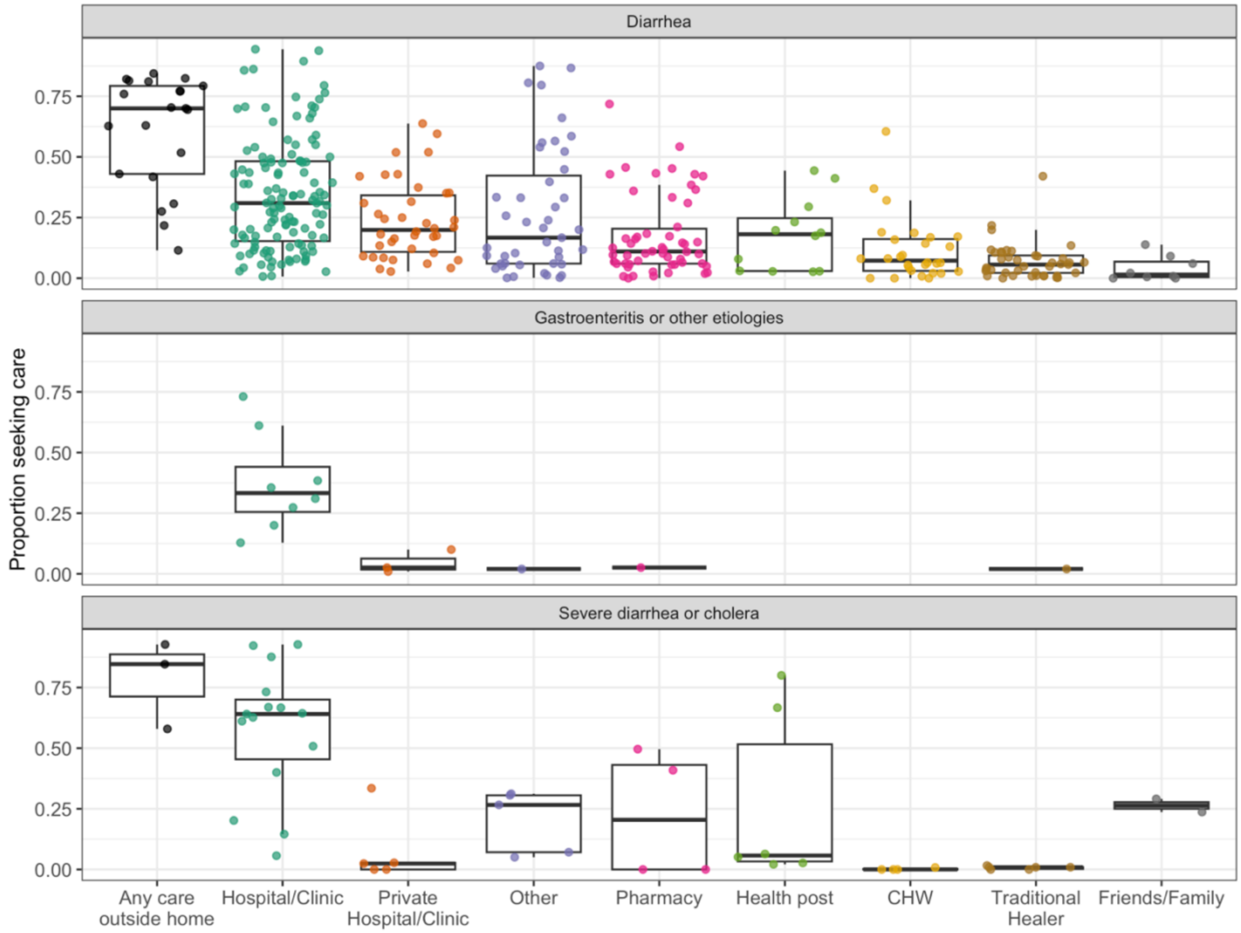

Figure S8. Proportion that sought care by source and observation characteristic for clinic-based studies

Proportion of respondents in clinic-based studies in LMICs that reported seeking care for diarrheal illness at all categories of care sources A) as their first source of care and B) prior to their current hospital or clinic visit. Each point represents an observation. Boxes represent the median and interquartile range (IQR) of the proportion for each group. Lines extend from the top and bottom of box to the largest proportion value no further than 1.5 * IQR from the box.

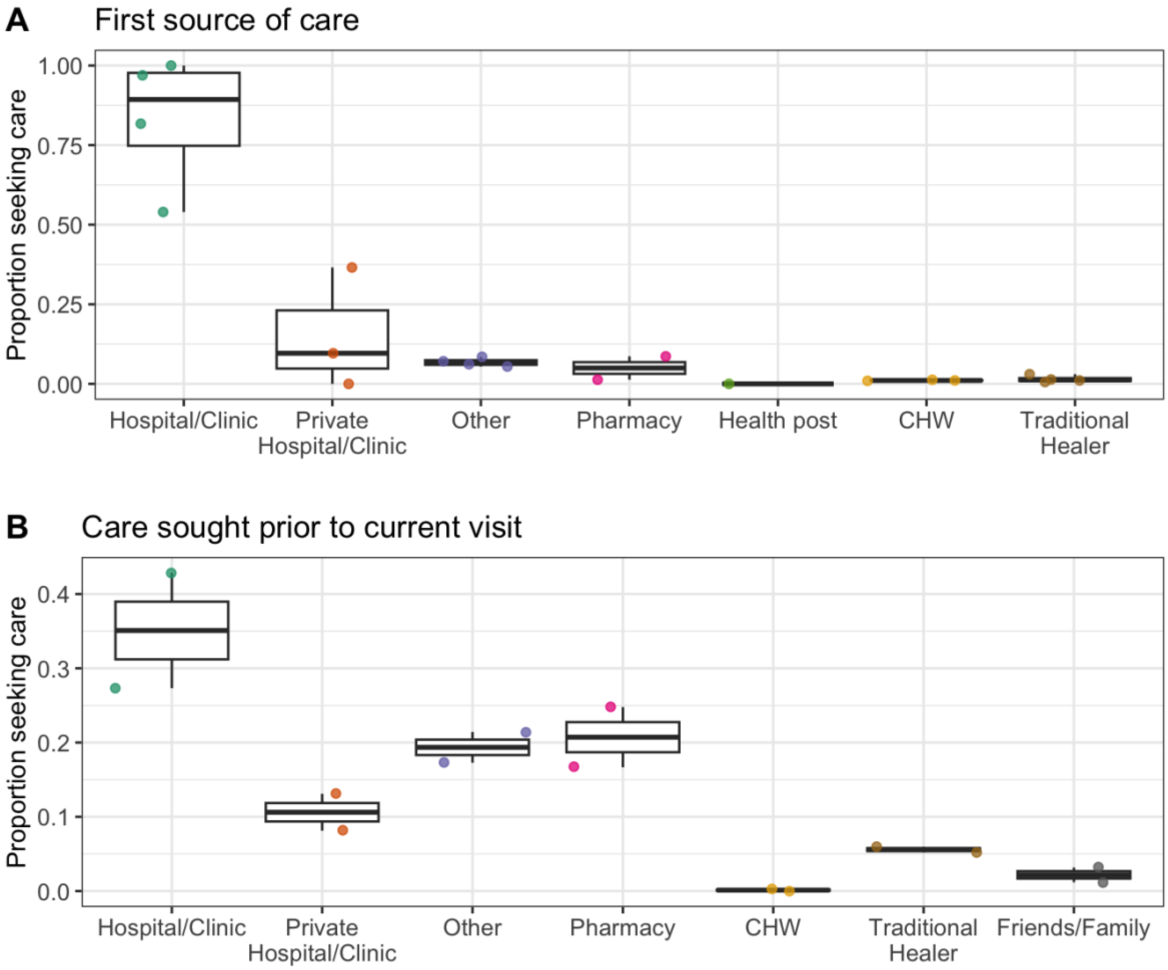

Figure S9. Forest plot of care seeking at a hospital or clinic in LMICs

Estimated proportion that sought care for an actual episode of diarrheal illness at a hospital or clinic across the 145 observations included in the random-effects meta-analysis.

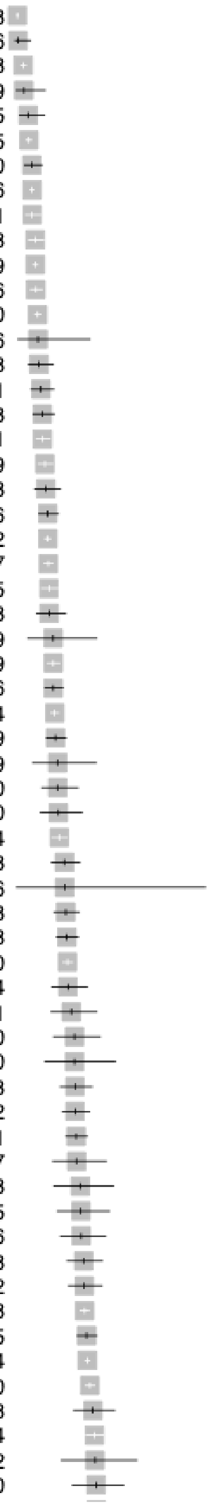

| Study | Events | Total | Proportion | 95%–CI |
|---|---|---|---|---|
| IND (125) Child, Diarrhea | 58 | 8038 | 0.01 | [0.01; 0.01] |
| BGD (26) Child, Diarrhea | 1 | 106 | 0.01 | [0.00; 0.05] |
| IND (134) Child, Diarrhea | 41 | 1508 | 0.03 | [0.02; 0.04] |
| IND (145) Combination, Diarrhea | 2 | 69 | 0.03 | [0.00; 0.10] |
| KEN (15) Combination, Diarrhea | 5 | 115 | 0.04 | [0.01; 0.10] |
| IND (85) Child, Diarrhea | 98 | 2145 | 0.05 | [0.04; 0.06] |
| BGD (31) Child, Diarrhea | 15 | 270 | 0.06 | [0.03; 0.09] |
| BGD (28) Combination, Diarrhea | 291 | 5156 | 0.06 | [0.05; 0.06] |
| GTM (14) Child, Severe diarrhea or cholera | 21 | 371 | 0.06 | [0.04; 0.09] |
| IND (24) Combination, Diarrhea | 29 | 428 | 0.07 | [0.05; 0.10] |
| BGD (92) Child, Diarrhea | 275 | 4049 | 0.07 | [0.06; 0.08] |
| UGA (178) Child, Diarrhea | 49 | 706 | 0.07 | [0.05; 0.09] |
| IND (100) Child, Diarrhea | 211 | 2800 | 0.08 | [0.07; 0.09] |
| IND (175) Self, Diarrhea | 2 | 26 | 0.08 | [0.01; 0.25] |
| KEN (15) Child, Diarrhea | 15 | 188 | 0.08 | [0.05; 0.13] |
| LBR (101) Child, Diarrhea | 20 | 231 | 0.09 | [0.05; 0.13] |
| PAK (163) Combination, Diarrhea | 26 | 283 | 0.09 | [0.06; 0.13] |
| BGD (116) Child, Diarrhea | 62 | 671 | 0.09 | [0.07; 0.12] |
| PAK (163) Child, Diarrhea | 63 | 629 | 0.10 | [0.08; 0.13] |
| GTM (17) Combination, Diarrhea | 22 | 213 | 0.10 | [0.07; 0.15] |
| GTM (17) Self, Diarrhea | 40 | 366 | 0.11 | [0.08; 0.15] |
| CMR (127) Child, Diarrhea | 367 | 3342 | 0.11 | [0.10; 0.12] |
| UGA (178) Child, Diarrhea | 185 | 1657 | 0.11 | [0.10; 0.13] |
| BRA (108) Combination, Diarrhea | 61 | 535 | 0.11 | [0.09; 0.14] |
| TZA (23) Child, Diarrhea | 21 | 183 | 0.11 | [0.07; 0.17] |
| LCA (64) Combination, Gastroenteritis or other etiologies | 5 | 39 | 0.13 | [0.04; 0.27] |
| MWI (36) Child, Diarrhea | 136 | 1059 | 0.13 | [0.11; 0.15] |
| MLI (171) Child, Diarrhea | 65 | 506 | 0.13 | [0.10; 0.16] |
| BGD (129) Child, Diarrhea | 333 | 2504 | 0.13 | [0.12; 0.15] |
| UGA (203) Child, Diarrhea | 61 | 449 | 0.14 | [0.11; 0.17] |
| KEN (123) Combination, Diarrhea | 7 | 49 | 0.14 | [0.06; 0.27] |
| UGA (203) Combination, Diarrhea | 20 | 140 | 0.14 | [0.09; 0.21] |
| GTM (14) Child, Severe diarrhea or cholera | 16 | 110 | 0.15 | [0.09; 0.23] |
| KEN (91) Child, Diarrhea | 121 | 814 | 0.15 | [0.12; 0.17] |
| MMR (32) Child, Diarrhea | 42 | 253 | 0.17 | [0.12; 0.22] |
| THA (93) Self, Diarrhea | 1 | 6 | 0.17 | [0.00; 0.64] |
| ZAF (192) Child, Diarrhea | 55 | 323 | 0.17 | [0.13; 0.22] |
| KEN (13) Child, Diarrhea | 78 | 448 | 0.17 | [0.14; 0.21] |
| ZMB (33) Child, Diarrhea | 392 | 2220 | 0.18 | [0.16; 0.19] |
| GTM (17) Self, Diarrhea | 31 | 174 | 0.18 | [0.12; 0.24] |
| GTM (17) Combination, Diarrhea | 21 | 111 | 0.19 | [0.12; 0.27] |
| GHA (12) Child, Diarrhea | 22 | 110 | 0.20 | [0.13; 0.29] |
| JAM (103) Combination, Gastroenteritis or other etiologies | 10 | 50 | 0.20 | [0.10; 0.34] |
| IND (176) Child, Severe diarrhea or cholera | 47 | 233 | 0.20 | [0.15; 0.26] |
| UGA (137) Child, Diarrhea | 65 | 322 | 0.20 | [0.16; 0.25] |
| CHN (102) Combination, Diarrhea | 105 | 511 | 0.21 | [0.17; 0.24] |
| ZAF (22) Combination, Diarrhea | 18 | 87 | 0.21 | [0.13; 0.31] |
| GHA (166) Combination, Diarrhea | 16 | 73 | 0.22 | [0.13; 0.33] |
| BGD (25) Child, Diarrhea | 21 | 95 | 0.22 | [0.14; 0.32] |
| MLI (171) Child, Diarrhea | 28 | 126 | 0.22 | [0.15; 0.30] |
| BGD (99) Self, Diarrhea | 48 | 208 | 0.23 | [0.18; 0.29] |
| NIC (9) Child, Diarrhea | 54 | 232 | 0.23 | [0.18; 0.29] |
| BGD (92) Child, Diarrhea | 762 | 3258 | 0.23 | [0.22; 0.25] |
| UGA (46) Child, Diarrhea | 165 | 685 | 0.24 | [0.21; 0.27] |
| KEN (131) Combination, Diarrhea | 3339 | 13724 | 0.24 | [0.24; 0.25] |
| PAK (192) Child, Diarrhea | 809 | 3210 | 0.25 | [0.24; 0.27] |
| GTM (17) Child, Diarrhea | 44 | 168 | 0.26 | [0.20; 0.34] |
| ETH (82) Child, Diarrhea | 242 | 904 | 0.27 | [0.24; 0.30] |
| HTI (141) Combination, Diarrhea | 14 | 52 | 0.27 | [0.16; 0.41] |
| AFG (72) Child, Diarrhea | 30 | 110 | 0.27 | [0.19; 0.37] |

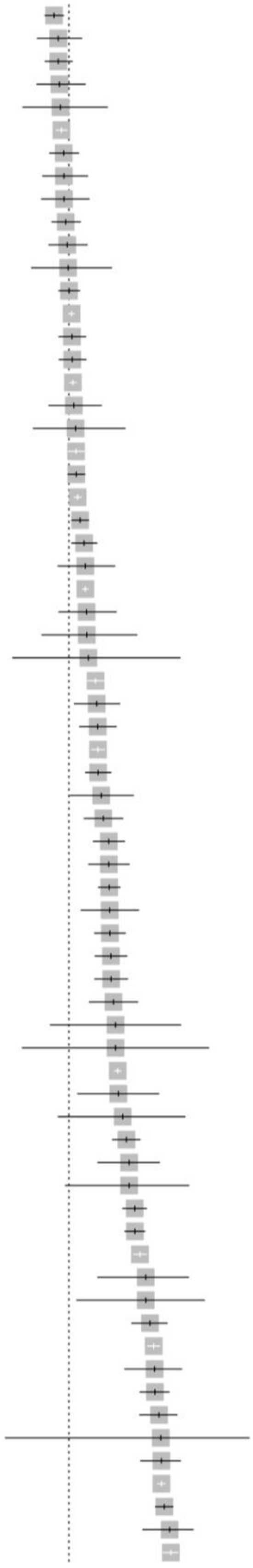

| | | | | |
|---|---|---|---|---|
| CUB (97) Combination, Gastroenteritis or other etiologies | 186 | 680 | 0.27 | [0.24; 0.31] |
| GHA (12) Child, Diarrhea | 37 | 128 | 0.29 | [0.21; 0.38] |
| MDG (96) Child, Diarrhea | 96 | 332 | 0.29 | [0.24; 0.34] |
| JOR (135) Combination, Diarrhea | 32 | 109 | 0.29 | [0.21; 0.39] |
| THA (167) Combination, Diarrhea | 11 | 37 | 0.30 | [0.16; 0.47] |
| PER (192) Child, Diarrhea | 632 | 2102 | 0.30 | [0.28; 0.32] |
| ETH (88) Child, Diarrhea | 94 | 304 | 0.31 | [0.26; 0.36] |
| GRD (118) Combination, Gastroenteritis or other etiologies | 40 | 129 | 0.31 | [0.23; 0.40] |
| ETH (3) Child, Diarrhea | 36 | 116 | 0.31 | [0.23; 0.40] |
| ZMB (89) Child, Diarrhea | 101 | 319 | 0.32 | [0.27; 0.37] |
| BRA (192) Child, Diarrhea | 58 | 180 | 0.32 | [0.25; 0.40] |
| PER (39) Child, Diarrhea | 14 | 43 | 0.33 | [0.19; 0.49] |
| ETH (168) Child, Diarrhea | 196 | 596 | 0.33 | [0.29; 0.37] |
| KEN (131) Child, Diarrhea | 2981 | 8817 | 0.34 | [0.33; 0.35] |
| KEN (113) Child, Diarrhea | 126 | 371 | 0.34 | [0.29; 0.39] |
| SEN (59) Child, Diarrhea | 125 | 367 | 0.34 | [0.29; 0.39] |
| NGA (30) Child, Diarrhea | 1858 | 5416 | 0.34 | [0.33; 0.36] |
| IND (128) Child, Diarrhea | 35 | 101 | 0.35 | [0.25; 0.45] |
| IND (94) Child, Diarrhea | 12 | 34 | 0.35 | [0.20; 0.54] |
| NPL (192) Child, Diarrhea | 382 | 1077 | 0.35 | [0.33; 0.38] |
| CHN (201) Combination, Gastroenteritis or other etiologies | 337 | 948 | 0.36 | [0.32; 0.39] |
| KEN (41) Child, Diarrhea | 1858 | 5154 | 0.36 | [0.35; 0.37] |
| IND (192) Child, Diarrhea | 360 | 974 | 0.37 | [0.34; 0.40] |
| CHN (146) Combination, Gastroenteritis or other etiologies | 171 | 445 | 0.38 | [0.34; 0.43] |
| GTM (17) Child, Diarrhea | 35 | 90 | 0.39 | [0.29; 0.50] |
| IDN (115) Combination, Diarrhea | 3140 | 8074 | 0.39 | [0.38; 0.40] |
| TZA (114) Child, Diarrhea | 35 | 89 | 0.39 | [0.29; 0.50] |
| ZAF (73) Child, Diarrhea | 13 | 33 | 0.39 | [0.23; 0.58] |
| KEN (142) Combination, Severe diarrhea or cholera | 4 | 10 | 0.40 | [0.12; 0.74] |
| BFA (111) Child, Diarrhea | 455 | 1067 | 0.43 | [0.40; 0.46] |
| MRT (195) Combination, Diarrhea | 62 | 144 | 0.43 | [0.35; 0.52] |
| PSE (2) Combination, Diarrhea | 96 | 221 | 0.43 | [0.37; 0.50] |
| KEN (53) Child, Diarrhea | 721 | 1656 | 0.44 | [0.41; 0.46] |
| ETH (27) Child, Diarrhea | 197 | 452 | 0.44 | [0.39; 0.48] |
| COD (154) Combination, Diarrhea | 34 | 76 | 0.45 | [0.33; 0.57] |
| PSE (2) Combination, Diarrhea | 91 | 200 | 0.46 | [0.38; 0.53] |
| IND (37) Child, Diarrhea | 144 | 303 | 0.48 | [0.42; 0.53] |
| CHN (49) Self, Diarrhea | 87 | 183 | 0.48 | [0.40; 0.55] |
| TZA (192) Child, Diarrhea | 297 | 623 | 0.48 | [0.44; 0.52] |
| IND (80) Child, Diarrhea | 44 | 92 | 0.48 | [0.37; 0.58] |
| MOZ (124) Child, Diarrhea | 150 | 313 | 0.48 | [0.42; 0.54] |
| NGA (19) Child, Diarrhea | 142 | 294 | 0.48 | [0.42; 0.54] |
| KEN (41) Child, Diarrhea | 133 | 275 | 0.48 | [0.42; 0.54] |
| UGA (158) Child, Diarrhea | 64 | 130 | 0.49 | [0.40; 0.58] |
| NGA (126) Child, Diarrhea | 9 | 18 | 0.50 | [0.26; 0.74] |
| ZAF (130) Child, Diarrhea | 4 | 8 | 0.50 | [0.16; 0.84] |
| KEN (41) Child, Severe diarrhea or cholera | 3314 | 6524 | 0.51 | [0.50; 0.52] |
| THA (109) Child, Diarrhea | 24 | 47 | 0.51 | [0.36; 0.66] |
| CHN (49) Child, Diarrhea | 10 | 19 | 0.53 | [0.29; 0.76] |
| KEN (113) Child, Diarrhea | 210 | 389 | 0.54 | [0.49; 0.59] |
| ETH (21) Child, Diarrhea | 44 | 80 | 0.55 | [0.43; 0.66] |
| ZAF (73) Child, Diarrhea | 11 | 20 | 0.55 | [0.32; 0.77] |
| MOZ (8) Child, Diarrhea | 291 | 510 | 0.57 | [0.53; 0.61] |
| BGD (86) Child, Diarrhea | 413 | 723 | 0.57 | [0.53; 0.61] |
| BGD (192) Child, Diarrhea | 985 | 1670 | 0.59 | [0.57; 0.61] |
| BGD (4) Child, Severe diarrhea or cholera | 22 | 36 | 0.61 | [0.43; 0.77] |
| GHA (165) Child, Gastroenteritis or other etiologies | 11 | 18 | 0.61 | [0.36; 0.83] |
| HTI (50) Child, Severe diarrhea or cholera | 141 | 225 | 0.63 | [0.56; 0.69] |
| HTI (50) Combination, Severe diarrhea or cholera | 1155 | 1803 | 0.64 | [0.62; 0.66] |
| HTI (69) Combination, Severe diarrhea or cholera | 56 | 87 | 0.64 | [0.53; 0.74] |
| ETH (197) Child, Diarrhea | 207 | 321 | 0.64 | [0.59; 0.70] |
| MWI (107) Child, Diarrhea | 128 | 194 | 0.66 | [0.59; 0.73] |
| KEN (142) Child, Severe diarrhea or cholera | 2 | 3 | 0.67 | [0.09; 0.99] |
| IND (48) Child, Diarrhea | 113 | 169 | 0.67 | [0.59; 0.74] |
| BGD (186) Combination, Severe diarrhea or cholera | 3154 | 4716 | 0.67 | [0.66; 0.68] |
| KEN (140) Child, Diarrhea | 566 | 833 | 0.68 | [0.65; 0.71] |
| KEN (202) Child, Diarrhea | 72 | 103 | 0.70 | [0.60; 0.79] |
| NER (112) Child, Diarrhea | 750 | 1066 | 0.70 | [0.68; 0.73] |

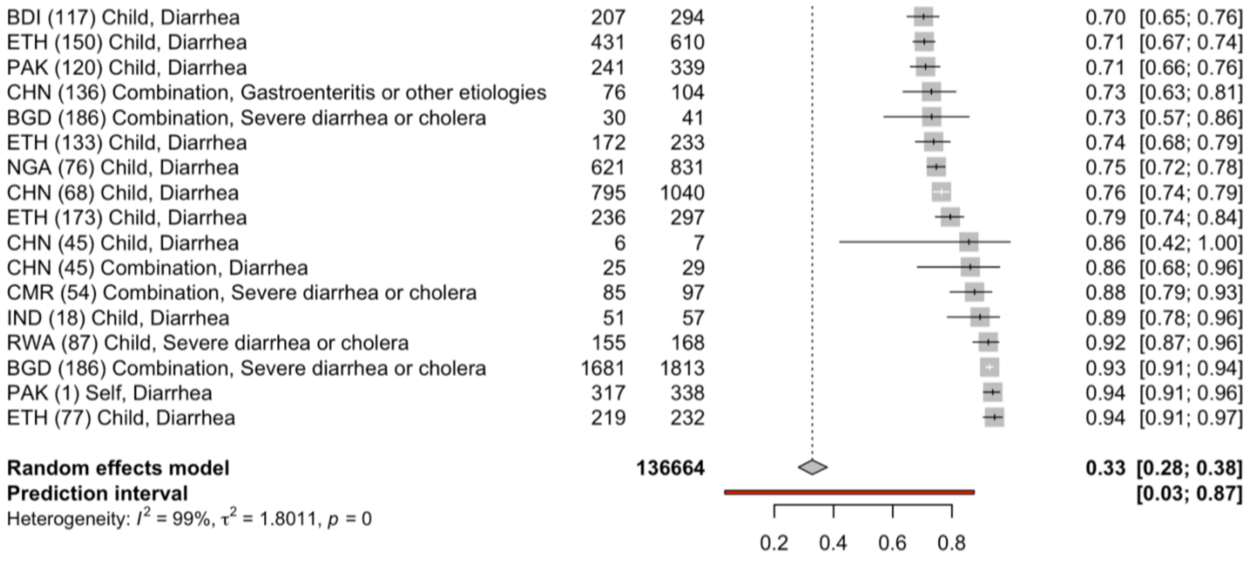
BDI (117) Child, Diarrhea 207 294 0.70 [0.65; 0.76]
ETH (150) Child, Diarrhea 431 610 0.71 [0.67; 0.74]
PAK (120) Child, Diarrhea 241 339 0.71 [0.66; 0.76]
CHN (136) Combination, Gastroenteritis or other etiologies 76 104 0.73 [0.63; 0.81]
BGD (186) Combination, Severe diarrhea or cholera 30 41 0.73 [0.57; 0.86]
ETH (133) Child, Diarrhea 172 233 0.74 [0.68; 0.79]
NGA (76) Child, Diarrhea 621 831 0.75 [0.72; 0.78]
CHN (68) Child, Diarrhea 795 1040 0.76 [0.74; 0.79]
ETH (173) Child, Diarrhea 236 297 0.79 [0.74; 0.84]
CHN (45) Child, Diarrhea 6 7 0.86 [0.42; 1.00]
CHN (45) Combination, Diarrhea 25 29 0.86 [0.68; 0.96]
CMR (54) Combination, Severe diarrhea or cholera 85 97 0.88 [0.79; 0.93]
IND (18) Child, Diarrhea 51 57 0.89 [0.78; 0.96]
RWA (87) Child, Severe diarrhea or cholera 155 168 0.92 [0.87; 0.96]
BGD (186) Combination, Severe diarrhea or cholera 1681 1813 0.93 [0.91; 0.94]
PAK (1) Self, Diarrhea 317 338 0.94 [0.91; 0.96]
ETH (77) Child, Diarrhea 219 232 0.94 [0.91; 0.97]
Random effects model 136664 0.33 [0.28; 0.38]
Prediction interval [0.03; 0.87]
Heterogeneity: $I^2$ = 99%, $\tau^2$ = 1.8011, $p$ = 0
0.2 0.4 0.6 0.8

Figure S10. Forest plot of hypothetical care seeking at a hospital or clinic in LMICs

Estimated proportion that would have sought care for a hypothetical episode of diarrheal illness at a hospital or clinic across the 29 observations included in the random-effects meta-analysis.

| Study | Events | Total | Proportion | 95%-CI |
|---|---|---|---|---|
| BGD (199) Self, Diarrhea | 16 | 148 | 0.11 | [0.06; 0.17] |
| BGD (199) Combination, Diarrhea | 330 | 2220 | 0.15 | [0.13; 0.16] |
| GHA (196) Combination, Diarrhea | 53 | 297 | 0.18 | [0.14; 0.23] |
| BGD (199) Child, Diarrhea | 22 | 111 | 0.20 | [0.13; 0.28] |
| GHA (196) Combination, Diarrhea | 29 | 122 | 0.24 | [0.17; 0.32] |
| THA (167) Combination, Diarrhea | 78 | 224 | 0.35 | [0.29; 0.41] |
| IRQ (156) Child, Diarrhea | 182 | 500 | 0.36 | [0.32; 0.41] |
| GHA (157) Child, Diarrhea | 3952 | 8715 | 0.45 | [0.44; 0.46] |
| GHA (12) Child, Diarrhea | 229 | 431 | 0.53 | [0.48; 0.58] |
| ETH (95) Child, Diarrhea | 951 | 1630 | 0.58 | [0.56; 0.61] |
| THA (167) Child, Diarrhea | 143 | 224 | 0.64 | [0.57; 0.70] |
| MWI (36) Combination, Diarrhea | 940 | 1403 | 0.67 | [0.64; 0.69] |
| IND (84) Child, Diarrhea | 475 | 670 | 0.71 | [0.67; 0.74] |
| GHA (12) Child, Diarrhea | 336 | 461 | 0.73 | [0.69; 0.77] |
| ETH (198) Combination, Diarrhea | 353 | 478 | 0.74 | [0.70; 0.78] |
| ETH (198) Combination, Diarrhea | 358 | 475 | 0.75 | [0.71; 0.79] |
| ETH (198) Child, Diarrhea | 213 | 275 | 0.77 | [0.72; 0.82] |
| KEN (142) Combination, Severe diarrhea or cholera | 330 | 425 | 0.78 | [0.73; 0.82] |
| LBN (70) Self, Severe diarrhea or cholera | 445 | 553 | 0.80 | [0.77; 0.84] |
| VNM (155) Child, Diarrhea | 321 | 396 | 0.81 | [0.77; 0.85] |
| ETH (198) Child, Diarrhea | 274 | 324 | 0.85 | [0.80; 0.88] |
| PAK (1) Self, Diarrhea | 286 | 338 | 0.85 | [0.80; 0.88] |
| ZAF (62) Child, Diarrhea | 155 | 182 | 0.85 | [0.79; 0.90] |
| PER (67) Child, Diarrhea | 1045 | 1200 | 0.87 | [0.85; 0.89] |
| CHN (45) Combination, Diarrhea | 212 | 243 | 0.87 | [0.82; 0.91] |
| CHN (45) Child, Diarrhea | 233 | 266 | 0.88 | [0.83; 0.91] |
| NER (112) Child, Diarrhea | 1806 | 1841 | 0.98 | [0.97; 0.99] |
| MOZ (43) Self, Severe diarrhea or cholera | 437 | 440 | 0.99 | [0.98; 1.00] |
| COD (159) Self, Severe diarrhea or cholera | 360 | 360 | 1.00 | [0.99; 1.00] |
| **Random effects model** | | **24952** | **0.72** | **[0.57; 0.83]** |
| **Prediction interval** | | | | **[0.06; 0.99]** |

Heterogeneity: $I^2$ = 99%, $\tau^2$ = 3.2002, $p$ = 0

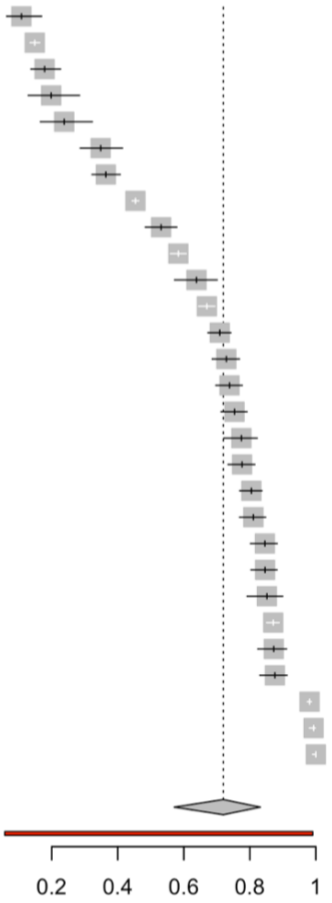

Figure S11. Forest plot of hypothetical care seeking at a hospital or clinic in HICs

Estimated proportion that sought care for an actual episode of diarrheal illness at a hospital or clinic across the 24 observations from high-income countries (HICs) included in the random-effects meta-analysis.

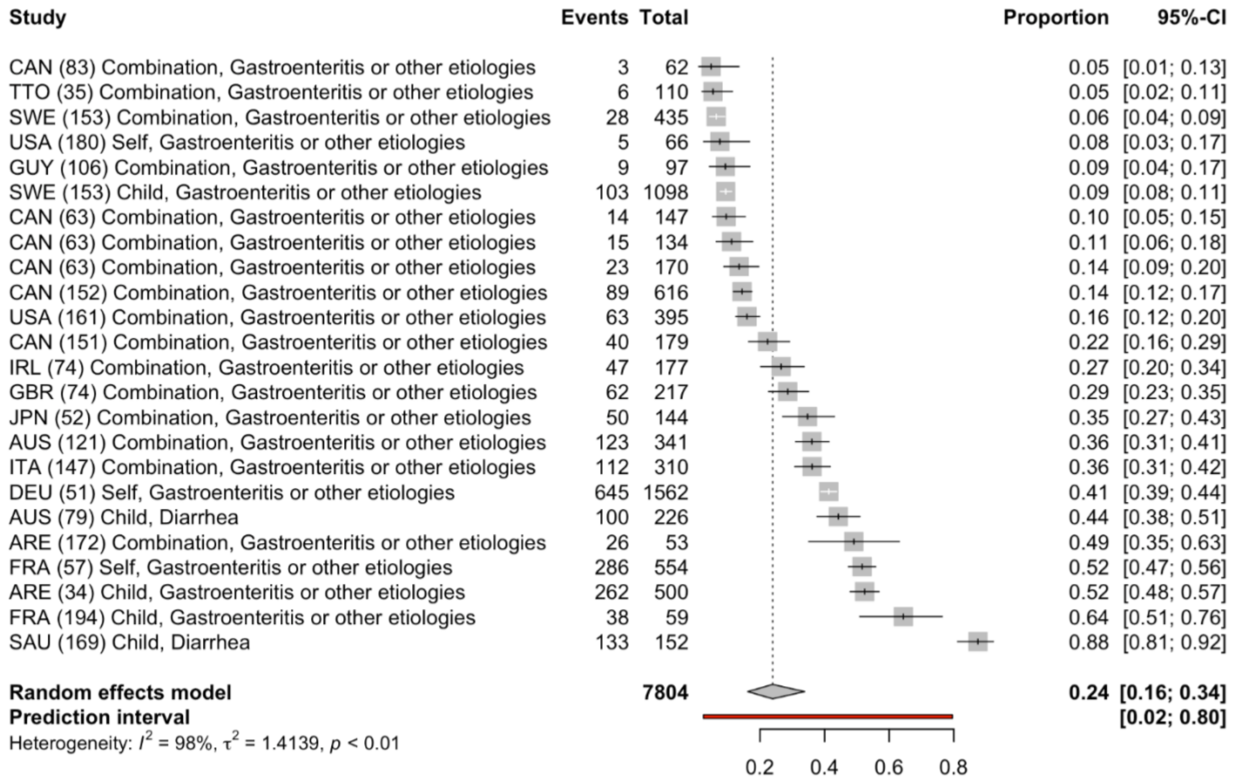

Figure S12. Forest plot of care seeking for general diarrhea in LMICs

Estimated proportion that sought care at a hospital or clinic across the 122 in LMICs studies that used a general diarrhea case definition.

| Study | Events | Total | Proportion | 95%-CI |
|---|---|---|---|---|
| IND (125) Child | 58 | 8038 | 0.01 | [0.01; 0.01] |
| BGD (26) Child | 1 | 106 | 0.01 | [0.00; 0.05] |
| IND (134) Child | 41 | 1508 | 0.03 | [0.02; 0.04] |
| IND (145) Combination | 2 | 69 | 0.03 | [0.00; 0.10] |
| KEN (15) Combination | 5 | 115 | 0.04 | [0.01; 0.10] |
| IND (85) Child | 98 | 2145 | 0.05 | [0.04; 0.06] |
| BGD (31) Child | 15 | 270 | 0.06 | [0.03; 0.09] |
| BGD (28) Combination | 291 | 5156 | 0.06 | [0.05; 0.06] |
| IND (24) Combination | 29 | 428 | 0.07 | [0.05; 0.10] |
| BGD (92) Child | 275 | 4049 | 0.07 | [0.06; 0.08] |
| UGA (178) Child | 49 | 706 | 0.07 | [0.05; 0.09] |
| IND (100) Child | 211 | 2800 | 0.08 | [0.07; 0.09] |
| IND (175) Self | 2 | 26 | 0.08 | [0.01; 0.25] |
| KEN (15) Child | 15 | 188 | 0.08 | [0.05; 0.13] |
| LBR (101) Child | 20 | 231 | 0.09 | [0.05; 0.13] |
| PAK (163) Combination | 26 | 283 | 0.09 | [0.06; 0.13] |
| BGD (116) Child | 62 | 671 | 0.09 | [0.07; 0.12] |
| PAK (163) Child | 63 | 629 | 0.10 | [0.08; 0.13] |
| GTM (17) Combination | 22 | 213 | 0.10 | [0.07; 0.15] |
| GTM (17) Self | 40 | 366 | 0.11 | [0.08; 0.15] |
| CMR (127) Child | 367 | 3342 | 0.11 | [0.10; 0.12] |
| UGA (178) Child | 185 | 1657 | 0.11 | [0.10; 0.13] |
| BRA (108) Combination | 61 | 535 | 0.11 | [0.09; 0.14] |
| TZA (23) Child | 21 | 183 | 0.11 | [0.07; 0.17] |
| MWI (36) Child | 136 | 1059 | 0.13 | [0.11; 0.15] |
| MLI (171) Child | 65 | 506 | 0.13 | [0.10; 0.16] |
| BGD (129) Child | 333 | 2504 | 0.13 | [0.12; 0.15] |
| UGA (203) Child | 61 | 449 | 0.14 | [0.11; 0.17] |
| KEN (123) Combination | 7 | 49 | 0.14 | [0.06; 0.27] |
| UGA (203) Combination | 20 | 140 | 0.14 | [0.09; 0.21] |
| KEN (91) Child | 121 | 814 | 0.15 | [0.12; 0.17] |
| MMR (32) Child | 42 | 253 | 0.17 | [0.12; 0.22] |
| THA (93) Self | 1 | 6 | 0.17 | [0.00; 0.64] |
| ZAF (192) Child | 55 | 323 | 0.17 | [0.13; 0.22] |
| KEN (13) Child | 78 | 448 | 0.17 | [0.14; 0.21] |
| ZMB (33) Child | 392 | 2220 | 0.18 | [0.16; 0.19] |
| GTM (17) Self | 31 | 174 | 0.18 | [0.12; 0.24] |
| GTM (17) Combination | 21 | 111 | 0.19 | [0.12; 0.27] |
| GHA (12) Child | 22 | 110 | 0.20 | [0.13; 0.29] |
| UGA (137) Child | 65 | 322 | 0.20 | [0.16; 0.25] |
| CHN (102) Combination | 105 | 511 | 0.21 | [0.17; 0.24] |
| ZAF (22) Combination | 18 | 87 | 0.21 | [0.13; 0.31] |
| GHA (166) Combination | 16 | 73 | 0.22 | [0.13; 0.33] |
| BGD (25) Child | 21 | 95 | 0.22 | [0.14; 0.32] |
| MLI (171) Child | 28 | 126 | 0.22 | [0.15; 0.30] |

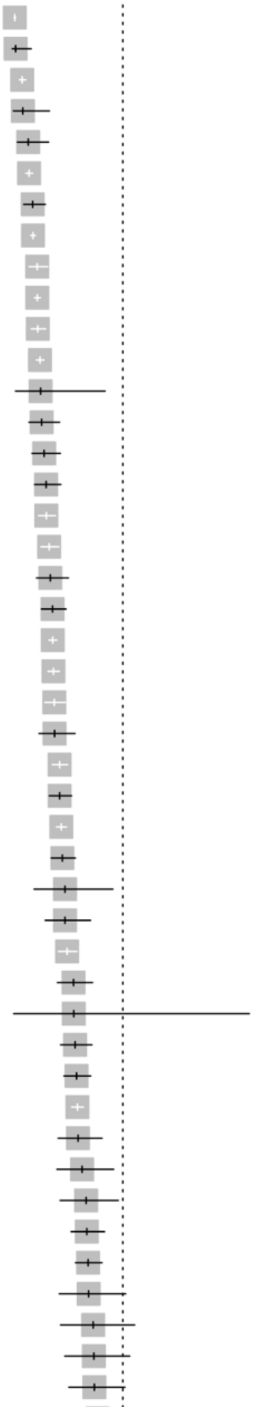

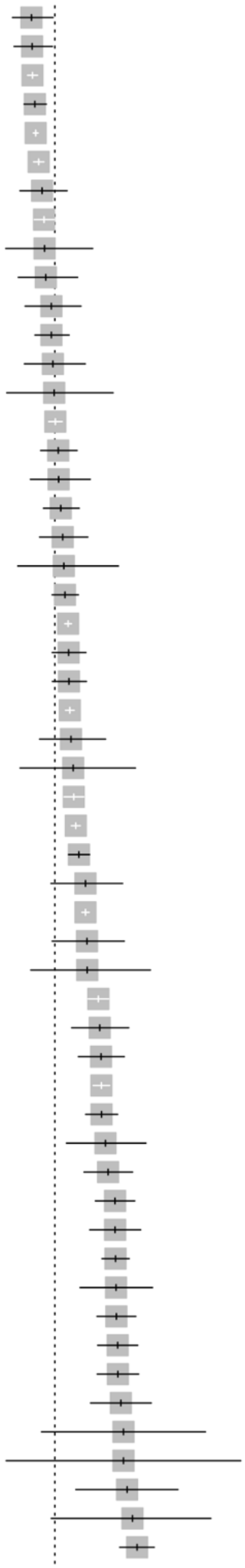

| | | | |
|---|---|---|---|
| BGD (99) Self | 48 | 208 | 0.23 [0.18; 0.29] |
| NIC (9) Child | 54 | 232 | 0.23 [0.18; 0.29] |
| BGD (92) Child | 762 | 3258 | 0.23 [0.22; 0.25] |
| UGA (46) Child | 165 | 685 | 0.24 [0.21; 0.27] |
| KEN (131) Combination | 3339 | 13724 | 0.24 [0.24; 0.25] |
| PAK (192) Child | 809 | 3210 | 0.25 [0.24; 0.27] |
| GTM (17) Child | 44 | 168 | 0.26 [0.20; 0.34] |
| ETH (82) Child | 242 | 904 | 0.27 [0.24; 0.30] |
| HTI (141) Combination | 14 | 52 | 0.27 [0.16; 0.41] |
| AFG (72) Child | 30 | 110 | 0.27 [0.19; 0.37] |
| GHA (12) Child | 37 | 128 | 0.29 [0.21; 0.38] |
| MDG (96) Child | 96 | 332 | 0.29 [0.24; 0.34] |
| JOR (135) Combination | 32 | 109 | 0.29 [0.21; 0.39] |
| THA (167) Combination | 11 | 37 | 0.30 [0.16; 0.47] |
| PER (192) Child | 632 | 2102 | 0.30 [0.28; 0.32] |
| ETH (88) Child | 94 | 304 | 0.31 [0.26; 0.36] |
| ETH (3) Child | 36 | 116 | 0.31 [0.23; 0.40] |
| ZMB (89) Child | 101 | 319 | 0.32 [0.27; 0.37] |
| BRA (192) Child | 58 | 180 | 0.32 [0.25; 0.40] |
| PER (39) Child | 14 | 43 | 0.33 [0.19; 0.49] |
| ETH (168) Child | 196 | 596 | 0.33 [0.29; 0.37] |
| KEN (131) Child | 2981 | 8817 | 0.34 [0.33; 0.35] |
| KEN (113) Child | 126 | 371 | 0.34 [0.29; 0.39] |
| SEN (59) Child | 125 | 367 | 0.34 [0.29; 0.39] |
| NGA (30) Child | 1858 | 5416 | 0.34 [0.33; 0.36] |
| IND (128) Child | 35 | 101 | 0.35 [0.25; 0.45] |
| IND (94) Child | 12 | 34 | 0.35 [0.20; 0.54] |
| NPL (192) Child | 382 | 1077 | 0.35 [0.33; 0.38] |
| KEN (41) Child | 1858 | 5154 | 0.36 [0.35; 0.37] |
| IND (192) Child | 360 | 974 | 0.37 [0.34; 0.40] |
| GTM (17) Child | 35 | 90 | 0.39 [0.29; 0.50] |
| IDN (115) Combination | 3140 | 8074 | 0.39 [0.38; 0.40] |
| TZA (114) Child | 35 | 89 | 0.39 [0.29; 0.50] |
| ZAF (73) Child | 13 | 33 | 0.39 [0.23; 0.58] |
| BFA (111) Child | 455 | 1067 | 0.43 [0.40; 0.46] |
| MRT (195) Combination | 62 | 144 | 0.43 [0.35; 0.52] |
| PSE (2) Combination | 96 | 221 | 0.43 [0.37; 0.50] |
| KEN (53) Child | 721 | 1656 | 0.44 [0.41; 0.46] |
| ETH (27) Child | 197 | 452 | 0.44 [0.39; 0.48] |
| COD (154) Combination | 34 | 76 | 0.45 [0.33; 0.57] |
| PSE (2) Combination | 91 | 200 | 0.46 [0.38; 0.53] |
| IND (37) Child | 144 | 303 | 0.48 [0.42; 0.53] |
| CHN (49) Self | 87 | 183 | 0.48 [0.40; 0.55] |
| TZA (192) Child | 297 | 623 | 0.48 [0.44; 0.52] |
| IND (80) Child | 44 | 92 | 0.48 [0.37; 0.58] |
| MOZ (124) Child | 150 | 313 | 0.48 [0.42; 0.54] |
| NGA (19) Child | 142 | 294 | 0.48 [0.42; 0.54] |
| KEN (41) Child | 133 | 275 | 0.48 [0.42; 0.54] |
| UGA (158) Child | 64 | 130 | 0.49 [0.40; 0.58] |
| NGA (126) Child | 9 | 18 | 0.50 [0.26; 0.74] |
| ZAF (130) Child | 4 | 8 | 0.50 [0.16; 0.84] |
| THA (109) Child | 24 | 47 | 0.51 [0.36; 0.66] |
| CHN (49) Child | 10 | 19 | 0.53 [0.29; 0.76] |
| KEN (113) Child | 210 | 389 | 0.54 [0.49; 0.59] |

| | | | | |
|---|---|---|---|---|
| ETH (21) Child | 44 | 80 | 0.55 | [0.43; 0.66] |
| ZAF (73) Child | 11 | 20 | 0.55 | [0.32; 0.77] |
| MOZ (8) Child | 291 | 510 | 0.57 | [0.53; 0.61] |
| BGD (86) Child | 413 | 723 | 0.57 | [0.53; 0.61] |
| BGD (192) Child | 985 | 1670 | 0.59 | [0.57; 0.61] |
| ETH (197) Child | 207 | 321 | 0.64 | [0.59; 0.70] |
| MWI (107) Child | 128 | 194 | 0.66 | [0.59; 0.73] |
| IND (48) Child | 113 | 169 | 0.67 | [0.59; 0.74] |
| KEN (140) Child | 566 | 833 | 0.68 | [0.65; 0.71] |
| KEN (202) Child | 72 | 103 | 0.70 | [0.60; 0.79] |
| NER (112) Child | 750 | 1066 | 0.70 | [0.68; 0.73] |
| BDI (117) Child | 207 | 294 | 0.70 | [0.65; 0.76] |
| ETH (150) Child | 431 | 610 | 0.71 | [0.67; 0.74] |
| PAK (120) Child | 241 | 339 | 0.71 | [0.66; 0.76] |
| ETH (133) Child | 172 | 233 | 0.74 | [0.68; 0.79] |
| NGA (76) Child | 621 | 831 | 0.75 | [0.72; 0.78] |
| CHN (68) Child | 795 | 1040 | 0.76 | [0.74; 0.79] |
| ETH (173) Child | 236 | 297 | 0.79 | [0.74; 0.84] |
| CHN (45) Child | 6 | 7 | 0.86 | [0.42; 1.00] |
| CHN (45) Combination | 25 | 29 | 0.86 | [0.68; 0.96] |
| IND (18) Child | 51 | 57 | 0.89 | [0.78; 0.96] |
| PAK (1) Self | 317 | 338 | 0.94 | [0.91; 0.96] |
| ETH (77) Child | 219 | 232 | 0.94 | [0.91; 0.97] |
| **Random effects model** | | **118014** | **0.30** | **[0.25; 0.35]** |
| **Prediction interval** | | | | **[0.03; 0.85]** |

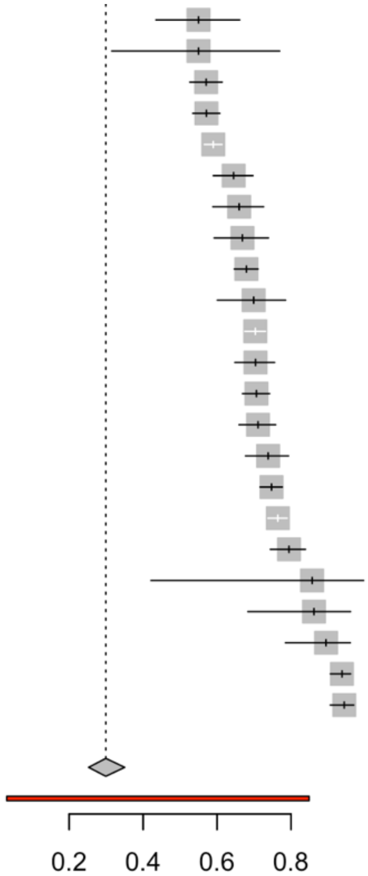


Heterogeneity: $I^2 = 99\%$, $\tau^2 = 1.6813$, $p = 0$

Figure S13. Forest plot of care seeking for severe diarrhea or cholera in LMICs

Estimated proportion that sought care at a hospital or clinic across the 15 studies in LMICs that used a severe diarrhea or cholera case definition, including deaths.

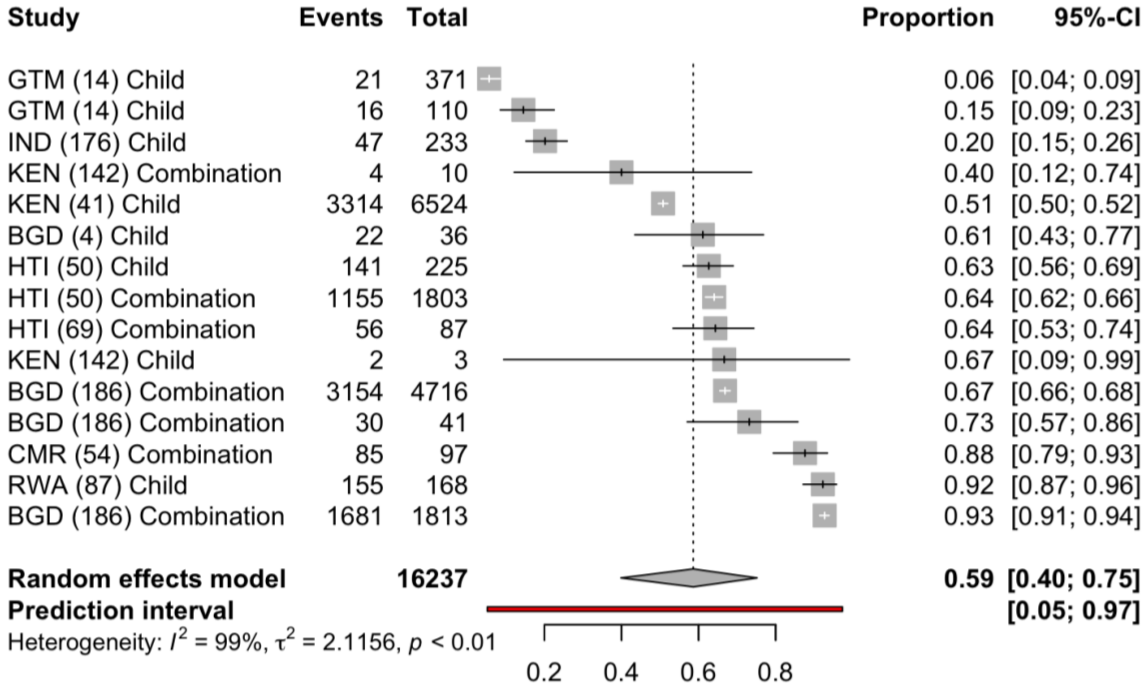


Figure S14. Forest plot of care seeking for gastroenteritis or other etiologies in LMICs

Estimated proportion that sought care at a hospital or clinic across the 8 studies in LMICs that used a gastroenteritis or other etiologies case definition.

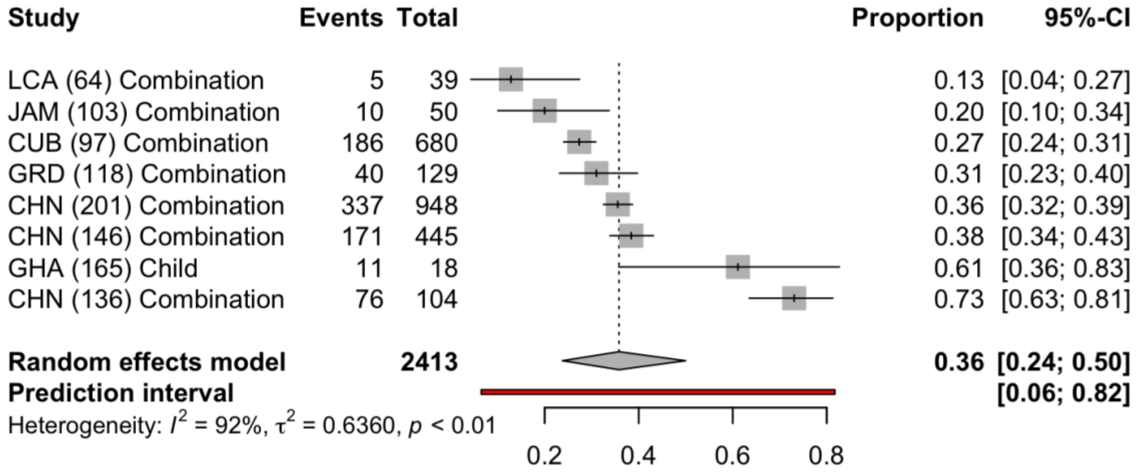

#### Supplementary Tables

##### Table S1. Standardized categories for sources of care and examples from studies

Standardized categories of sources of care sought outside the home and examples from included studies of options that fell into each of these categories.

| **Source of care** | **Example descriptions from papers** |
|---|---|
| Any care outside the home | ● Any non-specified source outside the home |
| Hospital or clinic | ● Doctor or licensed practitioner<br>● Hospital or clinic<br>● Qualified healthcare provider<br>● Cholera treatment center<br>● Free church-run clinic |
| Public hospital or clinic | ● Public hospital or clinic<br>● Public health center or facility<br>● District hospital<br>● Government clinic<br>● Government healthcare facility |
| Private hospital or clinic | ● Private hospital<br>● Private practitioner<br>● Private care provider |
| Pharmacy | ● Pharmacy or pharmacist<br>● Shop, market, or kiosk<br>● Drug or medicine seller<br>● Chemist |
| Health post | ● Health post<br>● Oral rehydration point<br>● Informal health clinic<br>● Community health center<br>● Dispensary |
| Community health worker | ● Community health worker<br>● Midwife<br>● Village health volunteer<br>● Community distributor |
| Traditional Healer | ● Traditional healer or provider<br>● Spiritual healer<br>● Religious leader<br>● Traditional medicine<br>● Bush doctor<br>● Homeopath |
| Friends/Family | ● Neighbor<br>● Friend<br>● Relative |
| Other | ● Unlicensed provider<br>● Informal help<br>● Village doctor<br>● Non-health facility<br>● Other |

Table S2. Univariate analyses of factors associated with variation in care seeking

Odds of seeking care for diarrhea at a hospital or clinic in a LMIC for each indicated variable category compared to the reference group, adjusting for no other variables. p-values are shown for the univariate mixed-effects models; n.s. indicates not significant ($p > 0.05$).

| **Variable** | **Category** | **Odds ratio** | **p-value** |
|---|---|---|---|
| Diarrhea case definition | Diarrhea | 1 [Reference] | |
| | Severe diarrhea or cholera | 3.26 (1.60 - 6.64) | < 0.01 |
| | Gastroenteritis or other etiologies | 1.3 (0.51 - 3.32) | n.s. |
| Study location type | Non-urban (Rural, Peri-Urban, and IDP or Refugee Camp) | 1 [Reference] | |
| | Urban | 1.08 (0.66 - 1.76) | n.s. |
| | Urban and non-urban | 1.12 (0.59 - 2.12) | n.s. |
| Multiple choices for types of care sought | No | 1 [Reference] | |
| | Yes | 1.00 (0.55 - 1.79) | n.s. |
| During or recently after an outbreak that causes diarrhea | No | 1 [Reference] | |
| | Yes | 3.15 (1.38 – 7.18) | < 0.01 |
| Survey recall period | 42-365 days or not reported | 1 [Reference] | |
| | 2-30 days | 0.44 (0.26 - 0.73) | < 0.01 |
| Relationship of diarrhea case to survey respondent | Child (of survey respondent) | 1 [Reference] | |
| | Combination (child, self, or other family member) | 0.91 (0.54 - 1.52) | n.s. |
| | Self (survey respondent) | 0.88 (0.30 - 2.57) | n.s. |
| Timing of care seeking | Care sought at any time | 1 [Reference] | |
| | First source sought | 0.46 (0.25 – 0.86) | < 0.05 |

Table S3. Tests for potential confounding with severe diarrhea or cholera case definition

Odds that an observation in a LMIC used severe diarrhea or cholera as the case definition instead of general diarrhea for each category compared to the reference group. p-values are shown for the univariate mixed-effects models; n.s. indicates not significant ($p > 0.05$).

| **Variable** | **Category** | **Odds ratio** | **p-value** |
|---|---|---|---|
| Multiple choices for types of care sought | No | 1 [Reference] | |
| | Yes | 0.93 (0.45 - 1.91) | n.s. |
| During or recently after an outbreak that causes diarrhea | No | 1 [Reference] | |
| | Yes | 3.52 (1.29 - 9.63) | < 0.05 |
| Survey recall period | 42-365 days or not reported | 1 [Reference] | |
| | 2-30 days | 0.47 (0.24 - 0.92) | < 0.05 |
| Relationship of diarrhea case to survey respondent | Child (of survey respondent) | 1 [Reference] | |
| | Combination (child, self, or other family member) | 1.40 (0.72 - 2.73) | n.s. |
| | Self (survey respondent) | 0.84 (0.24 - 2.93) | n.s. |
| Timing of care seeking | Care sought at any time | 1 [Reference] | |
| | First source sought | 1.11 (0.51 - 2.41) | n.s. |

Table S4. Sub-analyses of associations with outbreak context and recall period

Odds of seeking care at a hospital or clinic in a LMIC when a study describes an outbreak compared to when it does not among 15 studies with severe diarrhea or cholera case definition, and odds of seeking care when a study uses a recall period of 2-30 compared to longer recall periods among the 122 studies that used a general diarrhea case definition. p-values are shown for the univariate mixed-effects models; n.s. indicates not significant ($p > 0.05$).

| **Case definition for sub-analysis** | **Variable** | **Category** | **Odds ratio** | **p-value** |
|---|---|---|---|---|
| Severe diarrhea or cholera (n = 15) | During or recently after an outbreak that causes diarrhea | No | 1 [Reference] | |
| | | Yes | 1.52 (0.31 - 7.54) | n.s. |
| Diarrhea (n = 122) | Study recall period | 42-365 days or not reported | 1 [Reference] | |
| | | 2-30 days | 0.74 (0.39 - 1.43) | n.s. |
